# C allele of rs479200 of the host EGLN1 gene - a risk factor for severe COVID-19 (pilot study)

**DOI:** 10.1101/2022.03.11.22272214

**Authors:** Renuka Harit, Sajal De, Piyoosh Kumar Singh, Deepika Kashyap, Manish Kumar, Dibakar Sahu, Chander Prakash Yadav, Mradul Mohan, Vineeta Singh, Ram Singh Tomar, Kailash C Pandey, Kapil Vashisht

## Abstract

**Background:** Coronavirus disease-2019 (COVID-19) symptoms can range from asymptomatic, moderate to severe manifestations that result in an overall global case fatality rate of 2-7 %. While each variant has had it challenges, and some variants are more severe than others, risk factors of severe COVID-19 are still under investigation. In this context, the host genetic predisposition is also a crucial factor to investigate. In the present study, we investigated host genotypes of the SNP rs479200 of the host EGLN1 gene, previously implicated in high altitude pulmonary edema (HAPE), some of whose symptoms such as hypoxia profoundly overlap with severe COVID-19.

**Methods:** After informed consent, 158 RT-PCR confirmed COVID-19 patients (March 2020 to June 2021) were enrolled in the study. Based on their clinical manifestations, disease severity was categorized by the clinical team. Blood samples were drawn and DNA was extracted from the clot to infer different genotypes of the SNP rs479200 of the host EGLN1 gene. PCR-RFLP analysis of the SNP rs479200 (C > T) was performed with an amplicon size of 367 bp. Various genotypes (TT, TC and CC) were assigned based on the presence/absence of a restriction site (T/GTACA) for restriction enzyme *BsrGI*. Allele frequencies, Hardy-Weinberg Equilibrium (HWE) and multinomial logistic regression were performed using statistical tool SPSS version 23 (IBM).

**Findings:** We observed that the severe COVID-19 category was composed of comparatively younger patients with mean age (34.9±15.6), compared to asymptomatic and moderate categories whose mean age was 49.7±17.9 and 54.3±15.7, respectively. Preponderance of males and high heterozygosity (TC) was observed across the clinical categories. Notably, the frequency of C allele (0.664) was 2-fold higher than the T allele (0.336) in severe COVID-19 patients, whereas the allele frequencies were similar in asymptomatic and moderate category of COVID-19 patients. Multinomial logistic regression showed an association of genotypes with increasing clinical severity; odds ratio (adjusted OR-11.414 (2.564-50.812)) and (unadjusted OR-6.214 (1.84-20.99)) for the genotype CC in severe category of COVID-19. Interestingly, the TC genotype was also found to be positively associated with severe outcome (unadjusted OR-5.816 (1.489-22.709)), indicating association of C allele in imparting the risk of severe outcome.

**Interpretation:** The study provides strong evidence that the presence of C allele of SNP (rs479200) of the EGLN1 gene associates with severity in COVID-19 patients. Thus, the presence of C allele may be a risk factor for COVID-19 severity. This study opens new avenues towards risk assessment that include EGLN1 (rs479200) genotype testing and identifying patients with C allele who might be prioritized for critical care.

## Introduction

Severe acute respiratory syndrome coronavirus-2 (SARS-CoV-2) is the causal agent of coronavirus disease-2019 (COVID-19). The continuous evolution of the virus has generated different variants (Alpha, Beta, Gamma, Delta & Omicron) with their characteristic transmission and virulence profile. The clinical disease symptoms vary drastically, majority of the infections remain asymptomatic or moderate, often progressing to severe complications or mortality ranging from 2-7 % ^1^. Symptoms of severe COVID-19 pneumonia may include acute respiratory distress symptoms-hypoxia, dyspnea, hypocapnia or hypercapnia ^2^. However, the precise determinants of asymptomatic infections turning into severe disease are elusive. Interestingly, the COVID-19 hypoxia partially overlaps with high altitude pulmonary edema (HAPE) with some atypical features such as preserved lung compliance, intrapulmonary shunt and mild dyspnea. These features indicate towards loss of the homeostatic oxygen-sensing system which regulates oxygen uptake and systemic delivery. At molecular level, the mitochondrial proteins had been altered during SARS-CoV-2 infection leading to plausible disruptions in oxygen-sensingmechanisms ^3^. Several risk factors are being explored-both from the virus and host origin; these broadly include age, gender, BMI and pre-existing comorbidities ^4^. Thorough investigations of the host genetic predisposition is crucial as several genes have been reported to be associated with inflammation and immune responses and are considered as risk factors ^4^. Moreover, inter-individual variations in the effector or regulator genes might play role in progression to severe disease.

Focusing on the narrative of COVID-19 hypoxia, EGLN1 is one of the gene that has been comprehensively studied in context of HAPE hypoxia ^5^. The EGLN1 gene product- (2-oxoglutarate-dependent dioxygenase) regulates the hypoxia-inducible factor (HIF-α) *via* prolyl hydroxylation. Whereas in hypoxic conditions inactivation of EGLN1 leads to increased HIF-α and subsequent dimerization with HIF-β activate the hypoxia response elements (HRE) ^6,7^. The HRE response may include, but is not limited to alterations in metabolic, cellular and systemic responses inside the cells. Numerous studies in the literature have correlated the inter-individual variations of the EGLN1 gene (SNPs) in HAPE susceptibility ^5,8,9^, aerobic capacity ^10^, predisposition to preeclampsia ^11^ in different ethnicities. The TT genotype of the rs479200 SNP of the EGLN1 gene has been shown to associate with HAPE susceptibility ^5^. The EGLN1 gene on chromosome 1 translates into a 426 amino acid protein ^12^. Among the several genotypes of the EGLN1 gene, six of them (rs2472261, rs480902, rs479200, rs2808611, rs2153364, rs2491405) are present in the various ethnic populations of India ^13^. Specifically, rs479200 and rs480902 SNPs have been repeatedly implicated for significant associations with HAPE patients^5,14^. Mishra et al.^15^ have observed a multi-fold increase in EGLN1 expression in HAPE patients and significant association with the risk genotypes, their haplotypes and interacting genotypes. The arterial oxygen saturation (SaO_2_) levels were also found to be inversely correlated with pulmonary artery systolic pressure (PASP) and EGLN1 expression. SNPs from candidate genes have revealed significant differences of gene expression in HAPE patients and healthy controls-ACE (rs4309), HSP70 (rs1008438), SP-A2 (rs1965708), PAI-1 (rs1799889), NOS (rs199983) and EGLN1 (rs480902) etc. ^16^. The seven polymorphisms of EGLN1 gene-rs1538664, rs479200, rs2486729, rs2790879, rs480902, rs2486736 and rs973252; were found to be significantly different in HAPE patients and HAPE controls ^14^. A significant number of patients in severe COVID-19 mimic HAPE hypoxia ^17^.

The lung transcriptome of COVID-19 patient implicated interaction of viral proteins ORF8 and OS9 with EGLN1 and EGLN2 genes with potential role in disrupting the EGLN1-HIF oxygen sensor mechanism ^18,19^. It is worth noting that EGLN1 gene acts as a natural regulator of the HIF-α protein and hence corroborates the hypothesis that inter-individual variations in the EGLN1 gene might play role in the disease severity and subsequently risk assessment. Interestingly, Roxadustat-HIF inhibitor shown to reduce ACE-2 expression, thereby inhibiting entry and replication of SARS-CoV-2 inside lung epithelial cells, thus argued as potential therapeutic agents for COVID-19 ^20^. In the present study, categorized COVID-19 patients were analyzed for the SNP rs479200 of the EGLN1 gene and potential associations of the genotype with disease severity were investigated.

## Material and methods

### Sample collection and DNA isolation

Adult patients (age ≥ 18 years) with RT-PCR or Rapid Antigen Test positive for COVID-19 were included in the study. The patients were classified and defined by the clinical team as asymptomatic COVID-19 cases (no hospitalization); moderate COVID-19 infection (requiring oxygen support during hospitalization) and severe COVID-19 infection (required invasive/non-invasive ventilator support for > 24hrs) . Clotted blood samples of the COVID-19 patients were procured from the biorepository at ICMR-NIMR, Delhi and AIIMS, Raipur. The study was approved by the Institutional Ethics Committees (IECs), AIIMS, Raipur [1379/IEC-AIIMSRPR/2020] & ICMR-NIMR, Delhi [PHB/NIMR/EC/2020/145]. Nucleic acid extractions were performed at the COVID-19 testing facility at ICMR-NIMR, Delhi, using appropriate precautionary measures and personal protective equipment (PPE). DNA isolation from small fractions of the clotted blood was performed using DNeasy Blood and Tissue kit, Qiagen, Germany. Isolated DNA samples were quantified and purity was assessed using Quawell UV-Vis Spectrophotometer Q5000.

### Amplification for rs479200 and RFLP analysis

The EGLN1 gene sequence was retrieved from NCBI using Gene ID: 54583 for the egl-9 family hypoxia-inducible factor 1 [Homo sapiens (human)]. The reference SNP (rs479200) was identified from dbSNP Reference (rs) SNP report at the chromosomal position chr1:231408034 (GRCh38.p13) with alleles G>A / G>C. A 367 bp segment of the EGLN1 gene covering the SNP rs479200 was amplified using primers-FP-5‘*<u>CTCCCAGCACATCTGTGAAT</u>*3’ and RP-5‘*<u>TCGGATGGAAAGGTGGTAAAG</u>*3’. PCR amplification of the SNP region rs479200 with the above primers was performed using DreamTaq PCR Mastermix (2x) (Thermo Scientific, USA) in following steps-1) initial denaturation [95 °C / 3 minutes, 2) denaturation 95 °C / 30 seconds, 3) annealing 55 °C / 30 seconds, 4) extension 72 °C / 30 seconds, 5) steps 2 -4 for 34 cycles and 5) final extension 72°C / 5 minutes. The amplified products were visualized on 2% agarose gel and stored at 4 °C until further use.

For RFLP analysis, the 367 bp amplicon containing the rs479200 was cut with restriction enzyme *BsrGI* (T/GTACA) ^11^. The reaction comprised-10x Cut Smart Buffer, 5 U *BsrGI*-HF (New England Biolabs, USA), 100 ng of PCR amplicon containing rs479200 as template and incubated at 37 °C for 30 minutes. The digested products were visualized on 2% agarose gel to decipher the various genotypes of SNP rs4792000-homozygous (TT & CC), heterozygous (TC).

### Statistical analysis

The study data was fed in Microsoft Excel 365 and cross checked by two independent researchers. The statistical and association analysis was performed using the SPSS version 23 (IBM). The statistical analysis included the chi-square, genotype and allele frequency, Hardy-Weinberg analysis. Further the multinomial regression was employed to test the association of demographic and genetic factors.

## Results

Blood samples from 158 COVID-19 patients were tested by PCR-RFLP analysis to assess the genotypes of SNP rs479200 of the EGLN1 gene, using restriction enzyme *BsrGI* (T/GTACA). **Table 1** shows the demographic profiles of the patients included in the study, classified according to the disease severity. In our study, the mean age of the total number of participants was 45.9±18.3, however the mean age of severe COVID-19 patients was 34.9±15.6, contrary to the notion of prevalent severe complications in older patients. The mean age of COVID-19 patients in the asymptomatic and moderate disease was 49.7±17.9 and 54.3±15.7, respectively. In addition, preponderance of male COVID-19 patients was observed compared to females, across the clinical categories and also in a total number of participants, but it is unclear if this distribution is an attribute to social or biological causes.

**Table 1:** Demographic profiles of the COVID-19 patients classified according to disease severity.

|  |  | Asymptomatic |  | Moderate |  | Severe |  | Total |  |
| --- | --- | --- | --- | --- | --- | --- | --- | --- | --- |
|  |  | N | % | N | % | N | % | N | % |
| Age Mean ± SD | | 49.7 ± 17.9 | | 54.3 ± 15.7 | | 34.9 ± 15.6 | | 45.9 ± 18.3 | |
| Gender | Female | 9 | 23.70% | 11 | 17.70% | 18 | 31.00% | 38 | 22.75 |
|  | Male | 29 | 76.30% | 51 | 82.30% | 40 | 69.00% | 120 | 71.86 |

We then successfully amplified the EGLN1 gene segment (367 bp), flanking the SNP rs479200 (367 bp) from the DNA isolated from clotted blood of the COVID-19 patients **(Fig. 1)**.

**Figure 1:**
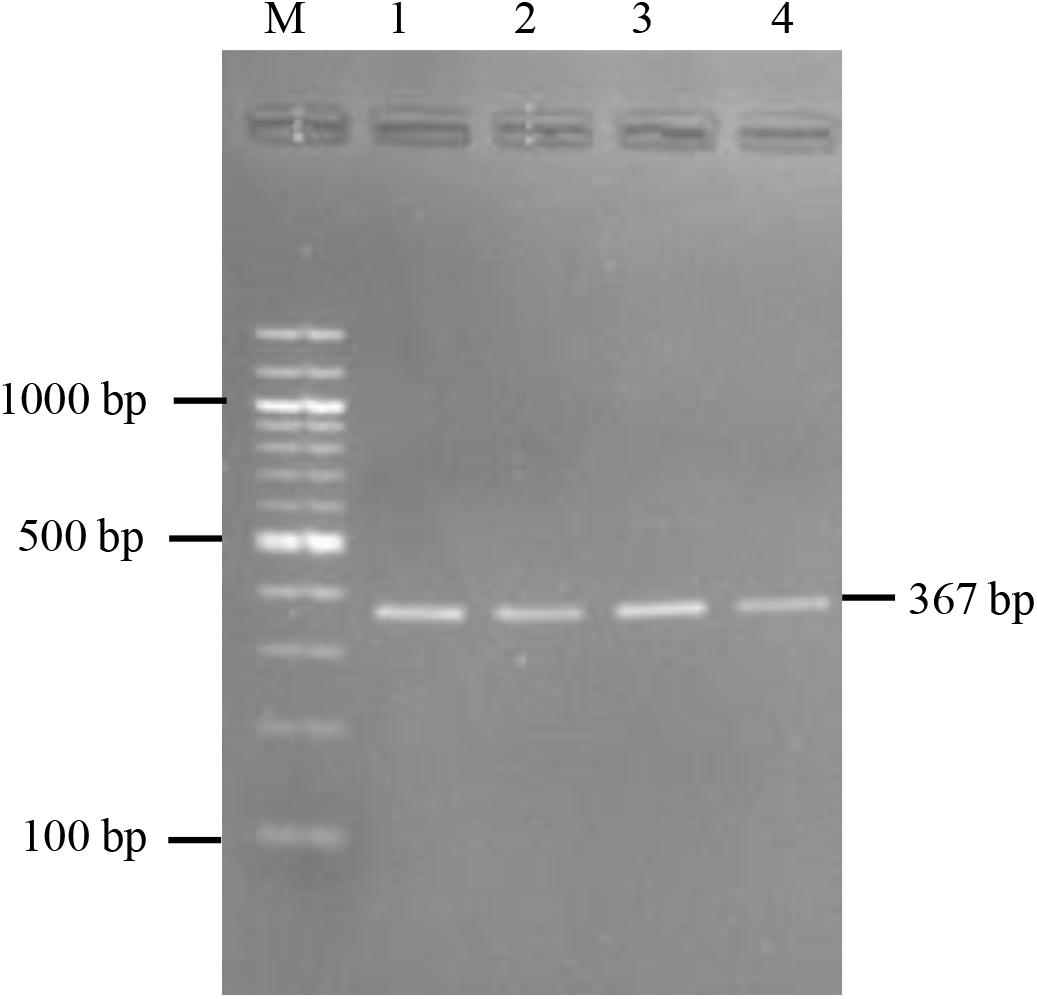
SNP rs479200 of the EGLN1 gene amplified from DNA isolated from clotted blood samples.

Further, PCR-RFLP analysis revealed the genotypes of the SNP rs479200 of the EGLN1 gene (**Fig. 2)**-*TT* genotype due to absence of ‘C’ at restriction site and hence no digestion (367 bp) by the restriction enzyme; *CC* genotype due to the presence of ‘C’ at restriction site leading to complete digestion (269 + 98 bp); *TC* genotype due to the presence of ‘C’ at restriction site, only in one allele and incomplete digestion conferring heterozygous allele (367 + 269 + 98 bp).

**Figure 2:**
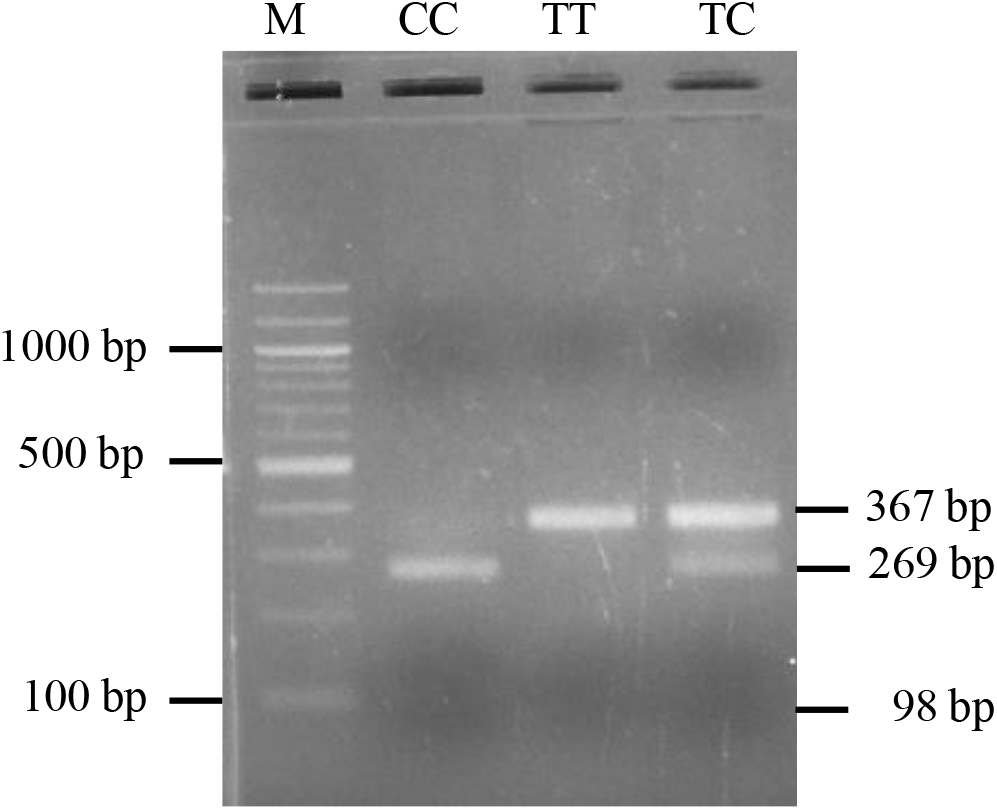
PCR-RFLP analysis of SNP rs479200 of EGLN1 gene with enzyme *BsrGI* (T/GTACA). The genotypes, allele frequencies of the EGLN1 gene rs479200 (T/C) and Hardy-Weinberg Equilibrium (HWE) of the study population are summarized in **Table 2**. The data showed high heterozygosity across the clinical categories and in the total number of study patients. It was observed that both the alleles (T & C) were uniformly present in the total study patients with a high proportion of ancestral T allele, except for the severe category patients. The frequency of ‘C’ allele (0.664) was observed to be 2-fold higher in proportion than the ‘T’ allele (0.336). The allele frequencies were in HWE status across the disease categories.

Multinomial logistic regression was performed in an adjusted and unadjusted model to test the association of genotype variables with clinical severity. Moderate and severe COVID-19 patients were compared with the asymptomatic patients, separately. In the unadjusted model the effects of genotype and disease severity were inferred, while the adjusted model, the genotypes were tested in isolation for their association with moderate and severe clinical outcomes. **Table 3** depicts the association analysis data, highlighting the association of genotype (CC) and genotype (TC) with severe outcome in COVID-19. The highest odds ratio (11.414 (2.564-50.812)) was observed with genotype (CC) in the adjusted model in asymptomatic vs severe COVID-19 subject’s comparison; however, the unadjusted model genotype (CC) shows the odds ratio of 6.214 (1.84-20.99). The heterozygous genotype (TC) was also found to be positively associated with disease severity (unadjusted OR-5.816 (1.489-22.709)). The genotypes did not demonstrate any association in the comparison scenario of moderate vs asymptomatic patients.

**Table 2:**
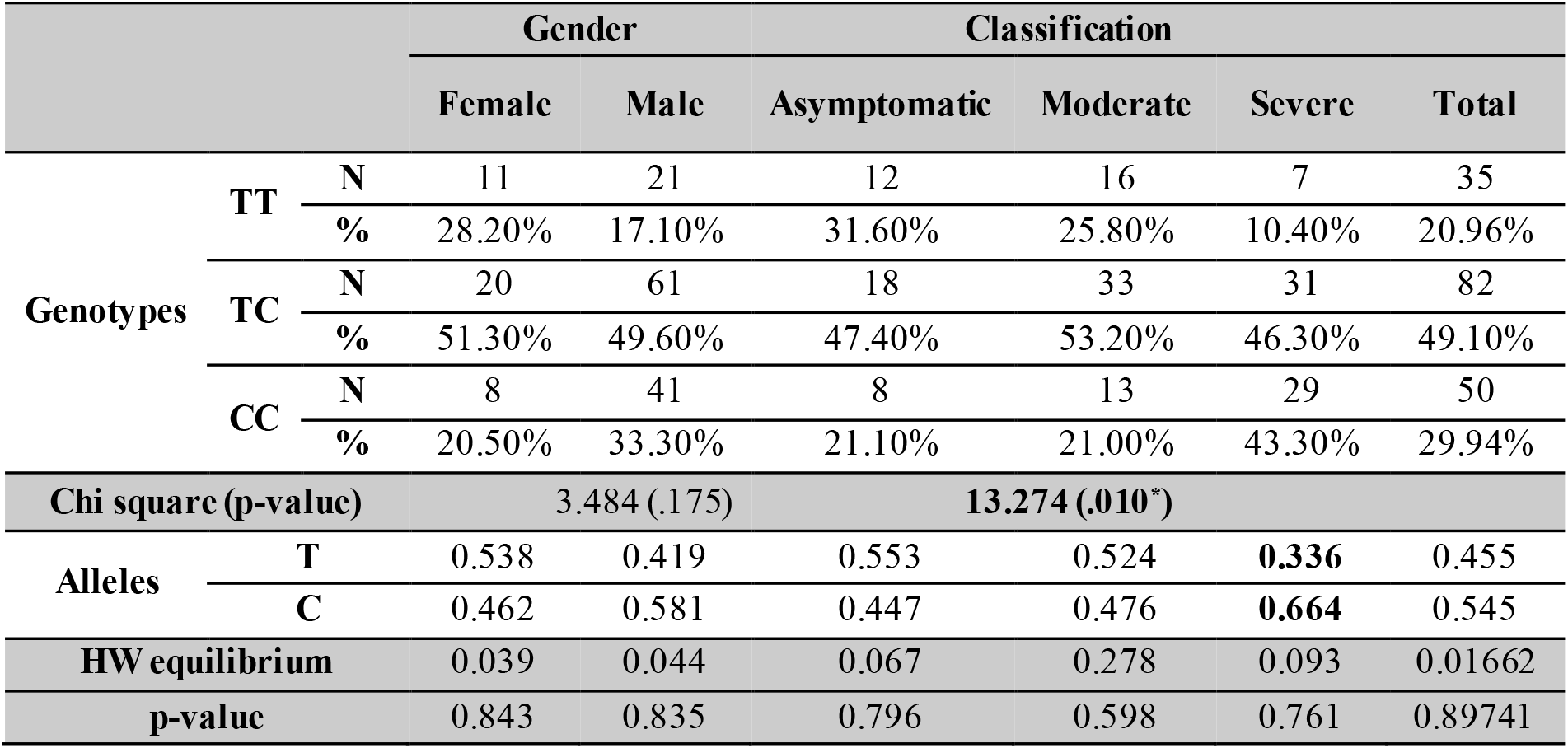
Genotypes, allele frequencies and Hardy–Weinberg Equilibrium (HWE) of the COVID-19 study patients.

**Table 3:** Multinomial logistic regression to infer the association of genotypes with disease severity.

| Variable (genotypes) |  | B- coefficient | Wald (Sig.) | Odds Ratio (CI) |
| --- | --- | --- | --- | --- |
| Asymptomatic Vs Moderate |  |  |  |  |
| Genotype (CC) | Adjusted | 0.077 | 0.016 (0.9) | 1.08 (0.326-3.576) |
|  | Unadjusted | 0.198 | 0.113 (0.737) | 1.219 (0.384-3.871) |
| Genotype (TC) | Adjusted | 0.199 | 0.162 (0.688) | 1.22 (0.463-3.215) |
|  | Unadjusted | 0.318 | 0.438 (0.508) | 1.375 (0.535-3.532) |
| Asymptomatic Vs Severe |  |  |  |  |
| Genotype (CC) | Adjusted | 2.435 | **10.213 (0.001)** | **11.414 (2.564-50.812)** |
|  | Unadjusted | 1.827 | **8.653 (0.003)** | **6.214 (1.84-20.99)** |
| Genotype (TC) | Adjusted | 1.761 | **6.417 (0.011)** | **5.816 (1.489-22.709)** |
|  | Unadjusted | 1.083 | **3.733 (0.053)** | **2.952 (0.984-8.854)** |

Reference genotype (TT)

## Discussion

Whilst there is a consensus that severe COVID-19 patients present acute respiratory distress, mimicking attributes of high altitude pulmonary edema (HAPE) ^2^. However, the similarities in the pathophysiology of severe COVID-19 and HAPE are debatable ^21^. Failure of respiratory homeostasis in acute respiratory distress syndrome (ARDS), severe COVID-19 and HAPE include variable hypoxia, dyspnoea, hypoxic pulmonary vasoconstriction (HPV), Ventilation/perfusion (V/Q) mismatch, pulmonary hypertension, pulmonary capillary pressure and reduced lung compliance ^1^. HIF-α is shown to be central to the oxygen-sensing mechanism recruited by cells in response to hypoxia, whose expression is regulated by 2 -oxoglutarate-dependent dioxygenase (2ODD-a product of EGLN1 gene); acting as a natural inhibitor of HIF-α. In normoxic conditions, a balanced proteasomal degradation of constitutively expressed HIF-1α induced by 2ODD abrogates the translocation of HIF-1α and HIF-β dimer to activate HRE transcription. However, in a hypoxic scenario the balance of 2ODD and HIF-1α skews towards overproduction of HIF-1α, leading to dimerization with HIF-β and transcriptional activation of HRE^22^. Higher levels of HIF-α have also been shown to induce higher expression of angiotensin-converting enzyme-2 (ACE-2) receptor on cell surface facilitating the viral entry in the case of SARS-CoV-2. Further, complicating the situation are the interindividual variations of the EGLN1 gene, that have been observed to be responsible for varying capabilities in coping with the anti-hypoxic responses.

The present study investigated the EGLN1 genotypes in asymptomatic, moderate and severe COVID-19 patients at the time of diagnosis. Particularly, SNP rs479200 of the EGLN1 gene that was previously found to be associated with HAPE susceptibility, aerobic capacity and predisposition to pre-eclampsia in multiple studies was explored further; where in SNP rs479200 (C > T) results in three distinct genotypes-homozygous (CC & TT) and heterozygous (TC), with C & T as major and minor alleles, respectively. Statistical analysis SNP rs479200 revealed that both the alleles are present in equal proportion in asymptomatic and moderate categories of COVID-19 patients, but overrepresentation of C allele (2-fold higher than T allele) in severe COVID-19 was peculiar. The CC genotype was significantly overrepresented in severe COVID-19 patients (p-value = 0.047). Multinomial logistic regression was performed to evaluate the association of different genotypes with disease severity, which estimated (OR-11.414 (2.564-50.812)- adjusted) and (OR- 6.214 (1.84-20.99)- unadjusted) for genotype CC, in asymptomatic vs severe categories of COVID-19. Moreover, the TC genotype was also found to be strongly associated with severe COVID-19, indicating the role of the C allele as a risk factor.

Transcription of the hypoxia response elements (HRE) in normoxic conditions is regulated and mediated by 2ODD- a gene product of EGLN1 gene, however in hypoxic conditions, inactive 2ODD cannot inhibit HIF-α and HIF-β dimerization leading to activation of HRE (**Fig. 3a, b**) Considering the genotype data of the EGLN1 gene, TT genotype is correlated with higher expression of EGLN1 gene resulting in overproduction of 2ODD ^5^ and conversely, the CC genotype results in lower expression of EGLN1 gene producing less 2ODD. Accordingly, Aggarwal et al. ^5^ previously showed that individuals residing at low altitudes with TT genotype do not respond well in hypoxic conditions succumbing to HAPE at high altitudes. Overproduction of 2ODD in the presence of T allele **(Fig. 3c)** results in minimal HIF-α levels, rendering the individuals hypo-responsive towards hypoxia. In contrast, the ancestral CC genotype in highlanders imparts a physiological adaptation to respond well in hypoxic conditions. Notably, C allele- the identified risk factor in severe COVID-19, producing inactive 2ODD in hypoxic conditions, accumulates high levels of HIF-α making them hyper-responsive towards hypoxia with activation of HRE **(Fig. 3d)**. The hyper-responsiveness towards hypoxia might explain the severe COVID-19 manifestations- pulmonary vascular alterations, overproduction of inflammatory cytokines, fluid accumulation in alveoli and the cytokine storm.

**Figure 3:**
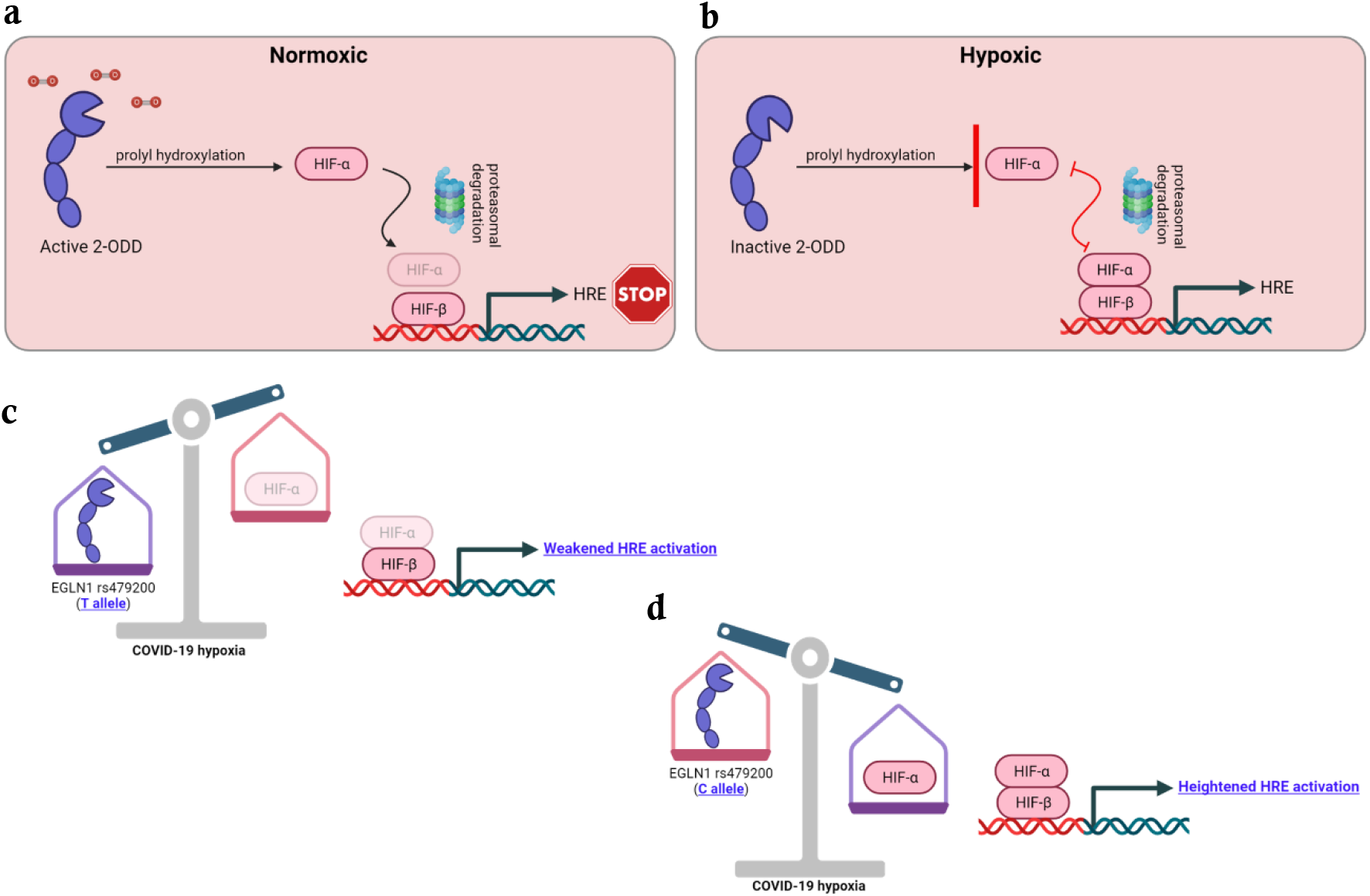
Schematic representation of interplay between HIF-α and 2ODD (product of EGLN1 gene), inhibiting activation of HRE in normoxic **(a)** and activation of HRE via dimerization with HIF-β in hypoxic **(b)** conditions. The postulated effect of T-allele **(c)** in COVID-19 hypoxia weakening the HRE activation and effect of C-allele **(d)** in the heightened HRE activation. (Created by BioRender trial version) (2ODD- 2-oxoglutarate-dependent dioxygenase; HIF-α- Hypoxia inducible factor- α; HIF-β- Hypoxia inducible factor- β; HRE- Hypoxic response elements.)

Thus, the hypothesis above postulates that individuals with severe COVID-19 symptoms and the C allele (risk factor), make them hyper-responsive in hypoxic conditions and progress towards worsening the respiratory hemodynamics and inflammatory pathways. To strengthen the hypothesis of C allele as a risk factor a larger cohort of categorized COVID-19 patients must be investigated in different settings. It is worth noting that deducing the genotype of SNP rs479200 can be easily accomplished in 4-6 hours’ time, equivalent to a COVID-19 RT-PCR test. Therefore, the presence of C allele as a risk factor in severe COVID-19 might serve as a feasible risk assessment prognostic marker and prioritization of limited medical infrastructure in view of COVID-19 waves, as0020witnessed globally002E

## Data Availability

All data produced in the present work are contained in the manuscript.

## Acknowledgement

This work was funded by intramural funding support from ICMR-NIMR, Delhi. We thank Drs. Amit Sharma and O. P. Singh for their critical feedback in the study. We thank the NIMR biorepository team members for procuring the COVID-19 infected patient samples during the first wave in India. The publication bears the NIMR publication committee no. RIC-11/2022. We thank Mr. Puneet Saini and Ramakant Pal from ICMR-NIMR, stores for timely procurement of reagents in unprecedented COVID-19 lockdown situation.

## Conflict of Interest

Authors declare no conflict of interest.

## Author contributions

Conceived and designed the experiments: KV, KCP, SD

Data generation and analysis: RH, PKS, DK, MK, DS, CPY, MM, VS, RST, KV, KCP

Data interpretation, manuscript writing and review: KV, KCP, PKS, CPY, VS, MM, SD

## Notes

### Competing Interest Statement

The authors have declared no competing interest.

### Author Declarations

The study was approved by the Institutional Ethics Committees (IECs), AIIMS, Raipur [1379/IEC-AIIMSRPR/2020] & ICMR-NIMR, Delhi [PHB/NIMR/EC/2020/145].

